# The demand for inpatient and ICU beds for COVID-19 in the US: lessons from Chinese cities

**DOI:** 10.1101/2020.03.09.20033241

**Authors:** Ruoran Li, Caitlin Rivers, Qi Tan, Megan B Murray, Eric Toner, Marc Lipsitch

## Abstract

**Background:** Sustained spread of SARS-CoV-2 has happened in major US cities. Capacity needs in Chinese cities could inform the planning of local healthcare resources.

**Methods:** We described the intensive care unit (ICU) and inpatient bed needs for confirmed COVID-19 patients in two Chinese cities (Wuhan and Guangzhou) from January 10 to February 29, 2020, and compared the timing of disease control measures in relation to the timing of SARS-CoV-2 community spread. We estimated the peak ICU bed needs in US cities if a Wuhan-like outbreak occurs.

**Results:** In Wuhan, strict disease control measures were implemented six weeks after sustained local transmission of SARS-CoV-2. Between January 10 and February 29, COVID-19 patients accounted for an average of 637 ICU patients and 3,454 serious inpatients on each day. During the epidemic peak, 19,425 patients (24.5 per 10,000 adults) were hospitalized, 9,689 (12.2 per 10,000 adults) were considered to be in serious condition, and 2,087 patients (2.6 per 10,000 adults) needed critical care per day. In Guangzhou, strict disease control measures were implemented within one week of case importation. Between January 24 and February 29, COVID-19 accounted for an average of 9 ICU patients and 20 inpatients on each day. During the epidemic peak, 15 patients were in critical condition, and 38 were classified as serious. If a Wuhan-like outbreak were to happen in a US city, the need for healthcare resources may be higher in cities with a higher prevalence of vulnerable populations.

**Conclusion:** Even after the lockdown of Wuhan on January 23, the number of seriously ill COVID-19 patients continued to rise, exceeding local hospitalization and ICU capacities for at least a month. Plans are urgently needed to mitigate the effect of COVID-19 outbreaks on the local healthcare system in US cities.

## Introduction

In the two months since the first report of four cases of atypical pneumonia in Wuhan, Hubei, China on December 27, 2019,^1^ the cumulative number of confirmed cases of COVID-19 in the city has risen to 49,122, with 2,195 deaths from the disease by the end of February, 2020.^2^ On January 23, Wuhan city shut down in response to the fast-evolving epidemic. All public transportation within, to and from the city was suspended, and residents were barred from leaving. An estimated nine million people remained in the city after the lockdown.^3^ Strict social distancing measures were also implemented, including the compulsory wearing of face-masks in public.

During the early phase of the response in Wuhan, the number of patients overwhelmed local fever clinics and hospitals designated to receive COVID-19 patients. The media reported a significant shortage of hospital beds, intensive care unit (ICU) beds, and other healthcare resources. By February 12, over eighteen thousand health care workers were sent to Wuhan from other parts of China to help with the coronavirus response.^4^ Forty eight hospitals (including two new hospitals built specifically for COVID-19 patients) and over 26,000 inpatient beds were designated for the isolation and treatment of patients with confirmed SARS-CoV2. Quarantine centers with over 13,000 total beds were also established to isolate confirmed patients with milder illnesses. By the end of February, the local government reported that “finally patients don’t need to wait for beds. Now the beds are waiting for patients.”^5^

With human-to-human transmission now established in other countries, mitigating the potential impact of COVID-19 on local healthcare systems is a top priority. A recent clinical study from China reported that 81% of patients in whom SARS-CoV-2 is detected experience mild disease, 14% severe disease and 5% critical disease.^1^ However, questions still remain as to the proportion of asymptomatic patients and the clinical course of the disease, preventing accurate prediction of hospitalization and ICU needs using transmission models.

Here, we describe the ICU and hospitalization needs for COVID-19 in two Chinese cities: Wuhan, the epicenter of China’s outbreak, and Guangzhou, a Chinese metropolis that experienced early importation of cases. As with all Chinese cities, strict social distancing measures and contact tracing and quarantine protocols were implemented since late January in Guangzhou, which resulted in much smaller outbreak size than Wuhan. Describing and comparing the resource needs in both cities may serve as benchmarks to help other large metropolises in the world prepare for potential outbreaks.

## Methods

We extracted and estimated confirmed COVID-19 case counts for severe and critical cases from Wuhan and Guangzhou from situation updates from Chinese national and local health commissions. We extracted the number of designated COVID-19 beds and hospitalizations from the Wuhan Municipal Health Commission website.

A confirmed COVID-19 case was considered severe if the patient experienced at least one of the following: dyspnea, respiratory frequency ≥30/minute, blood oxygen saturation ≤93%, arterial blood oxygen partial pressure (PaO2) to oxygen concentration (FiO2) ratio <300mmHg, and/or a pneumonia patient showing significant progression of lesions infiltrating >50% of the lung field on chest imaging within 24-48 hours. A confirmed patient was considered to be critical if he/she experienced respiratory failure demanding invasive and/or non-invasive ventilation for respiratory support, septic shock, and/or with multiple organ dysfunction/failure demanding intensive care.^6,7^ These definitions have been more detailed with revisions of the Chinese diagnostic and treatment guidelines. In this study, we used the term serious patients to describe severe and critical patients collectively. We estimated the number of prevalent severe and critical cases cross-sectionally per day - allowing for the fact that patients could move in and out of these categories over the course of their disease.

We extracted Wuhan city and Hubei province COVID-19 data between January 10 and February 29, including the numbers of confirmed cases, new cures, new deaths, severe cases, critical cases, serious cases (a sum of severe and critical cases), cumulative cures, cumulative deaths, cumulative confirmed cases, and currently confirmed cases (cumulative confirmed cases - deaths - cures). If official sources did not have data for variables on some dates, we calculated the number of cases based on the relationships between variables. Because Wuhan did not systematically report the number of severe and critical cases, we estimated these numbers by assuming that the proportions of serious and critical cases out of all currently confirmed cases was the same in Wuhan as in the rest of Hubei. For the dates when it was not possible to estimate the severe and critical case counts using the above methods (January 18, 25, and 27), we assumed the number of severe and critical cases on those dates were the same as reported for the previous day.

For Guangzhou, we extracted the city’s case count on the number of confirmed, severe, clinical, and cured cases and deaths for each day between January 24 and February 29.

### Statistical analysis

We summed the total patient-days under critical and/or severe condition to estimate the total ICU-days and serious-inpatient-days. We plotted the raw number of patients in critical and severe conditions and patients hospitalized on each day for Wuhan and Guangzhou, and estimated the proportion of hospitalization and ICU admission per 10,000 adults based on the assumption that there were 9 million people present in Wuhan during the lockdown,^3^ of whom 88.16% were age 15 or above (2010 census for cities in Hubei province), and 14.9 million present in Guangzhou of whom 82.82% were age 15 or above (Guangdong statistical bureau). We then projected the number of patients who have severe and critical COVID-19 disease at the peak of a Wuhan-like outbreak in the 30 most populous US cities by assuming that the effect of age and comorbidity on patient outcomes would be the same as their effect on COVID-19 mortality as derived from case reports from China until February 11.^8^ Specifically, we estimated the stratum-specific critical care rate in Wuhan by assuming that the risk factor for being in critical care is the same as that for death (age and comorbidities, e.g. hypertension).^8^ We estimated the probability of being in critical condition at the peak of the epidemic in each age and hypertension stratum using the COVID-19 mortality rate ratios for age and hypertension^8^ and the proportion of Wuhan population in each stratum. The hypertension prevalence in adults in Wuhan was estimated as 25.7%,^9^ and the proportion of the population aged over 65 years 14.1%.^10^ We then applied these stratum-specific critical care rate to the population structures in US cities based on the crude hypertension prevalence in adults in 2017^11^ and the proportion of adult population over 65 years of age in these cities.^12^

## Results

In Wuhan, COVID-19 accounted for a total of 32,486 ICU-days and 176,136 serious-inpatient-days between January 10 and February 29 (**Figure 1**), an average of 637 ICU patients and 3,454 serious inpatients on each day over that 51-day period. During the peak of the epidemic from mid to late February, a maximum of 19,425 patients (24.5 per 10,000 adults) were hospitalized, 9,689 patients (12.2 per 10,000 adults) were considered to be in “serious” condition, and 2,087 patients (2.6 per 10,000 adults) needed critical care per day.

**Figure 1.**
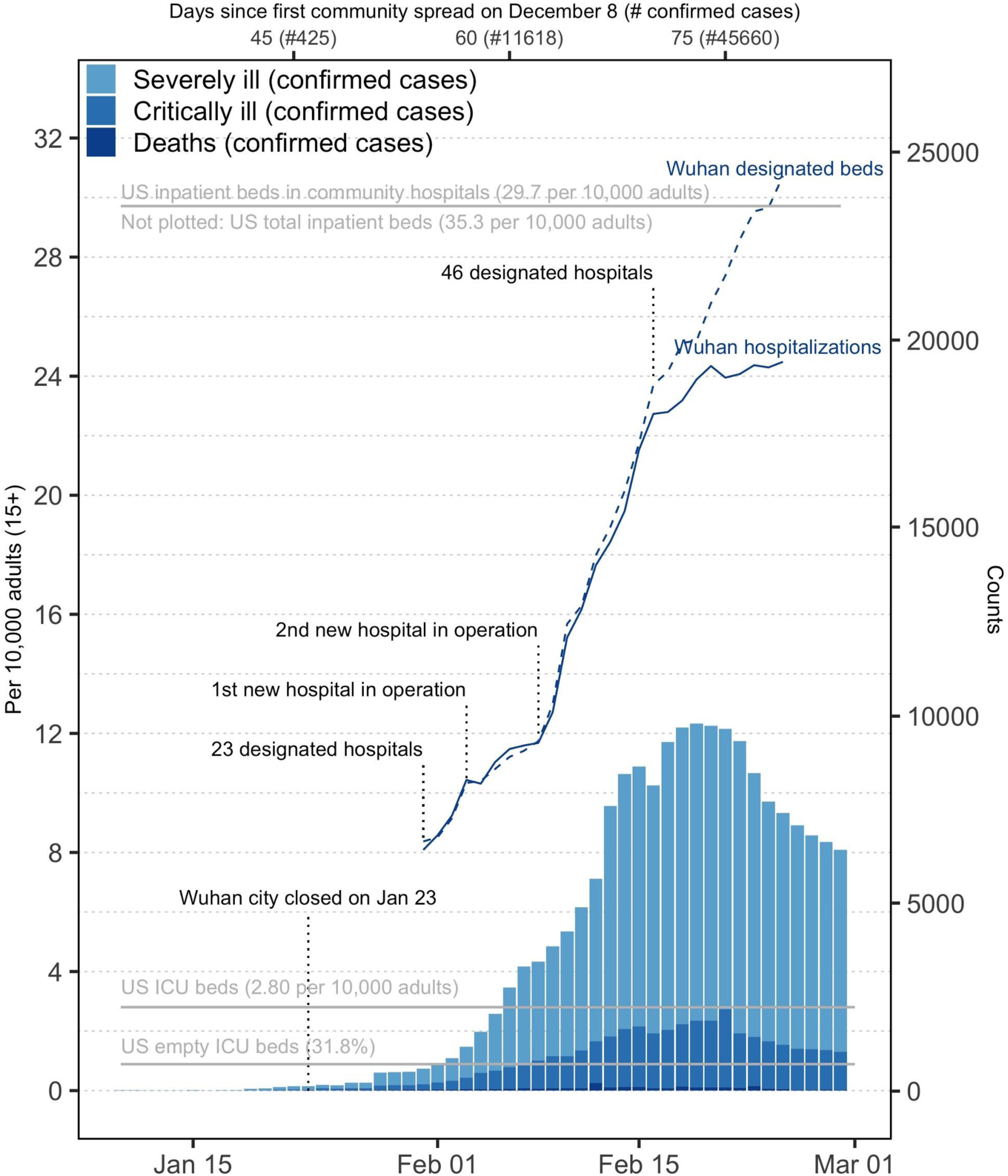
Burden of serious COVID-19 disease in Wuhan. US data sources: US ICU beds;^19^ US empty ICU beds;^20^ US inpatient beds in community hospitals;^21^ US population structure.^22^ The total number of US inpatient beds is 35.3 per 10,000 adults^23^.

In Guangzhou, COVID-19 accounted for a total of 318 ICU-days and 724 inpatient-days between January 24 and February 29 (**Figure 2**), an average of 9 ICU patients and 20 inpatients during that 37-day period. During the peak of the epidemic (early February), 15 patients were in critical condition, while 38 were hospitalized and classified as serious. Unlike Wuhan, where patients with mild COVID-19 disease were isolated in quarantine centers and not in designated hospitals, all confirmed patients in Guangzhou were hospitalized until cure. The maximum number of hospitalizations in Guangzhou on any day was 271 patients.

**Figure 2.**
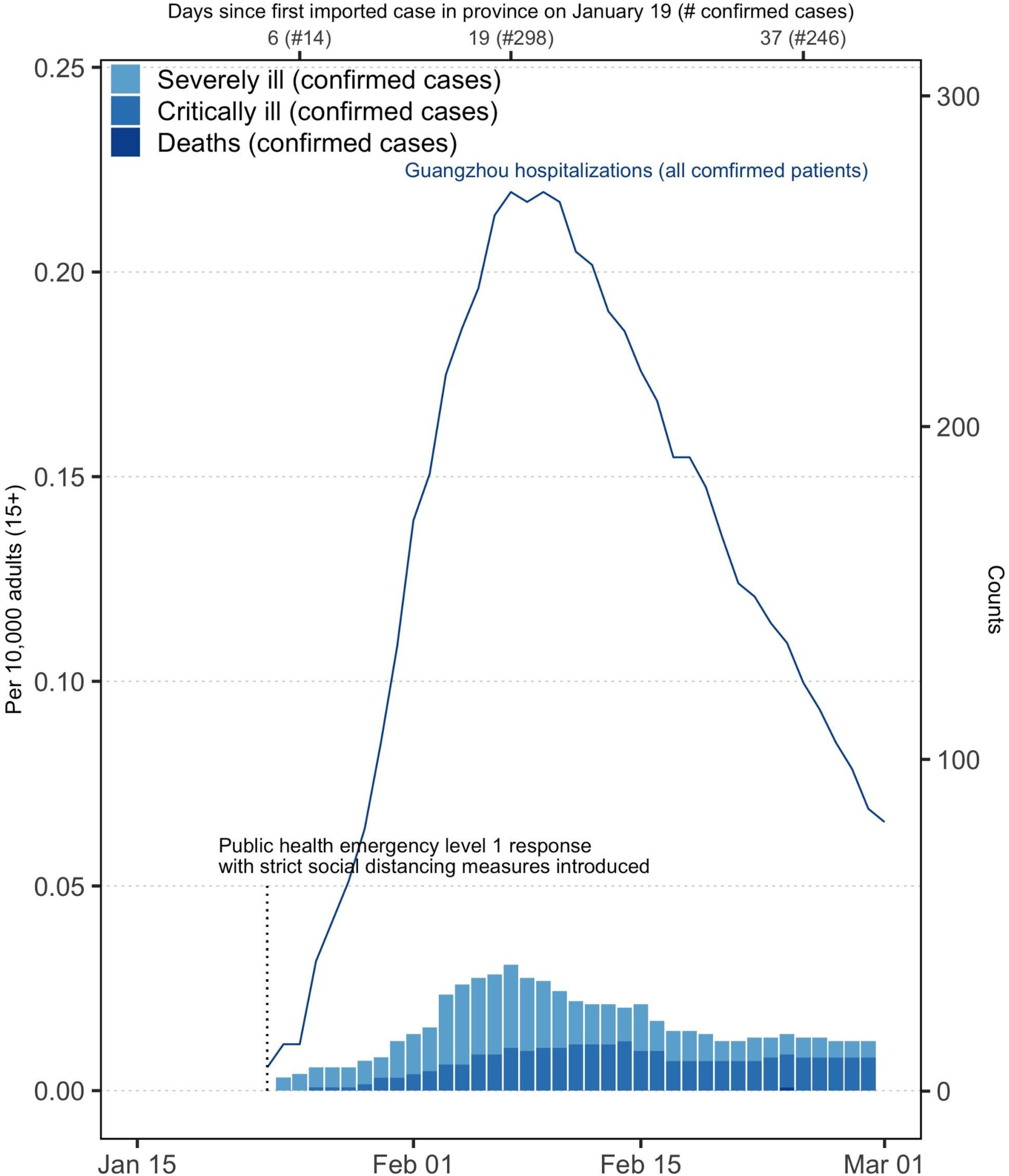
Burden of serious COVID-19 disease in Guangzhou.

The projected number of prevalent critically ill patients at the peak of a Wuhan-like outbreak in US cities ranges from 2.1 to 4.0 per 10,000 adults when we took into account of the difference in age distribution (**Figure 3, top**), and from 2.6 to 4.9 per 10,000 adults when we took into account of the differences in comorbidity (hypertension) prevalence (**Figure 3, bottom**).

**Figure 3.**
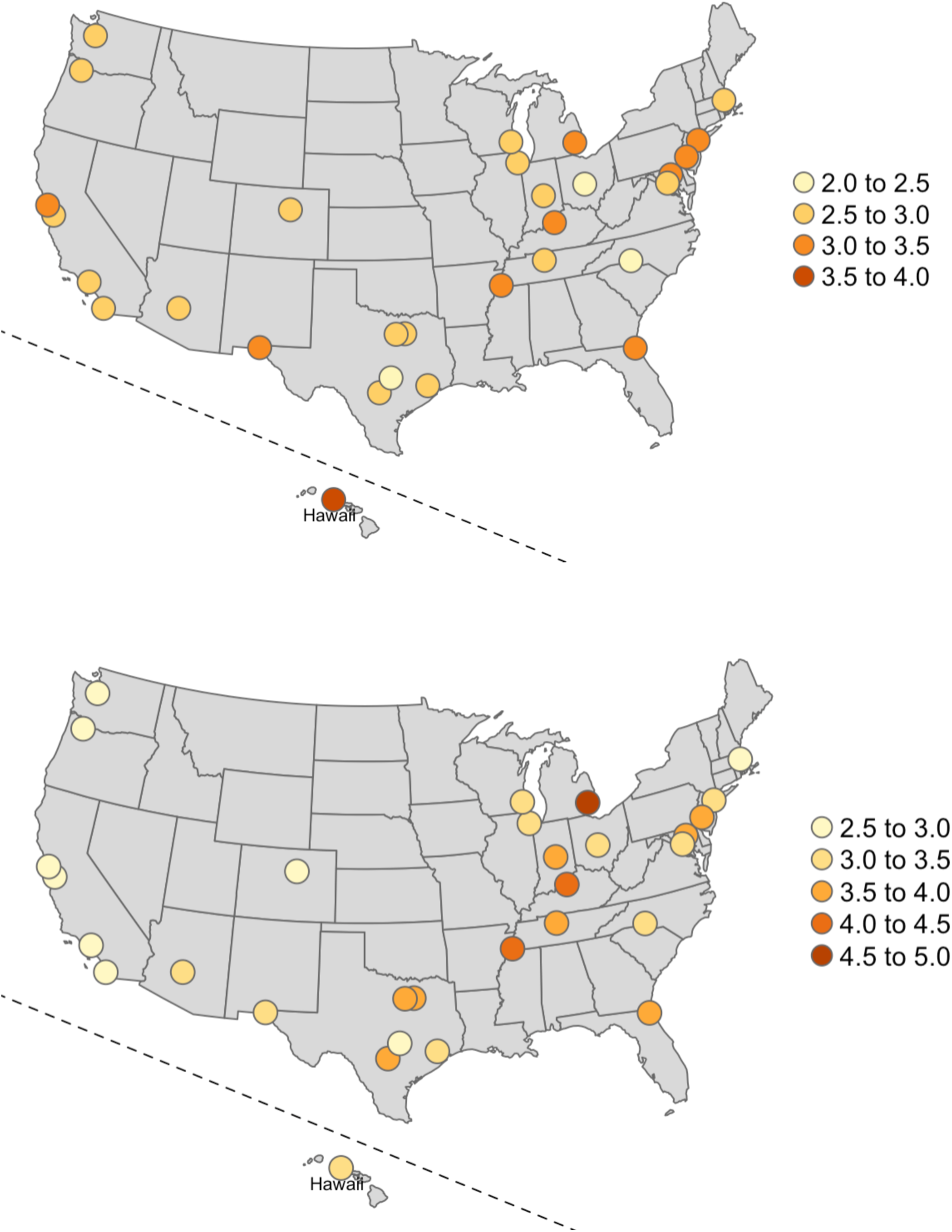
Estimate number of critically ill patients at the peak of a Wuhan-like outbreak in cities in the US, per 10,000 adults. Top: taking into account of the proportion of population over 65 years of age Bottom: taking into account of the proportion of population with hypertension Wuhan: 2.6 per 10,000 adults were critically ill at the peak of the COVID-19 epidemic, with a crude hypertension prevalence of 25.7% among adults (rate ratio for critical illness=6.9), and 15.9% adults over 64 years of age (rate ratio for critical illness=7.2).

## Discussion

Even after the lockdown of Wuhan on January 23, the number of seriously ill COVID-19 patients continued to rise, exceeding local hospitalization and ICU capacities for at least a month. During the peak of the Wuhan epidemic in February, nearly 20,000 COVID-19 patients were hospitalized simultaneously, with 10,000 in severe or critical conditions. If a Wuhan-like outbreak were to take place in a US city, even with strong social distancing and contact tracing protocols as strict as the Wuhan lockdown, hospitalization and ICU needs from COVID-19 patients alone may exceed current capacity. The need for healthcare resources may be higher in some US cities where there is a higher prevalence of vulnerable populations (age and comorbidity) than in Wuhan.

Exceeding healthcare capacity may increase the community spread of SARS-CoV-2. In Wuhan, home isolation and quarantine were used in the early phase of the epidemic to alleviate the demand in healthcare resources. However, because of the exponential increase of the number of patients who developed serious illness but could not be hospitalized due to capped capacity, secondary transmission in the community continued as patients and their household contacts moved between hospitals seeking care.

Exceeding healthcare capacity may also lead to decreased quality of care, such as not being able to get access to a ventilator, which would lead to an increased case fatality ratio. By the end of February, Wuhan’s case fatality ratio was 4.5%, 3.2% for the rest of Hubei province, and for the rest of China, where healthcare capacity was not exceeded due to strong social distancing and contact quarantine measures in the early phase of the epidemic (such as Guangzhou), 0.8%.^13^

In both Wuhan and Guangzhou, the lockdowns did not lead to immediate downturns in the demand for hospitalization or the number of serious cases; rather, the peak in these measures occurred approximately a month after the lockdown in Wuhan, and two weeks after the lockdown in Guangzhou. This delay reflects the potentially long time from infection to severe and critical conditions as many COVID-19 patients who eventually require ICU care present initially as having only mild symptoms,^14^ and even longer time to discharge or death,^15^ resulting in the accumulation of hospitalized cases long after the downturns in the community spread. In Wuhan, the longer delay may also reflect the ongoing transmission after the lockdown described above, which itself resulted from the overloading of the healthcare system.

This study has several limitations. We relied on officially reported statistics, which may not represent the change of actual case counts over time, but rather a reflection of testing and hospitalization capacity. The trend in Wuhan of the number of serious cases and hospitalizations is thus not reflective of actual need, but rather the trend in maximum capacity of the Wuhan system in diagnosis and treatment. We are therefore more confident of the hospitalization and serious case counts in Wuhan after mid-February, and in Guangzhou, where excess capacities in diagnosis and treatment were reported based on both official and unofficial sources. In addition, our projection of the ICU bed needs in US cities does not take into account scenarios where local transmission may differ from that of Wuhan.

The contact rate in Wuhan during the early phase of the epidemic may have been much higher than what we expect to occur in US cities because of the increased number of social contacts that occurred in Wuhan due to the Lunar New Year celebrations. If social distancing measures are effectively implemented early in US cities, the growth of the epidemic may be delayed. But it is also possible that US cities may not be able to implement the extreme social distancing measures that were put into place later in the epidemic in Wuhan. Therefore, the actual number of hospital and ICU beds that will be needed over the course of a COVID-19 outbreak in a US city is impossible to estimate precisely. Our estimated capacity needs based on a “Wuhan-like” outbreak could be a benchmark for what healthcare providers would expect to see during the first three months of a local COVID-19 epidemic.

Historical evidence has shown that in 1918, US cities which imposed nonpharmaceutical interventions early in the epidemic course and maintained their interventions over a long period had lower peaks and fewer total cases of pandemic influenza than those which waited.^16,17^ Our comparison of Wuhan and Guangzhou, although it is only two cities, dramatically illustrates the same relationship of early intervention leading to lower epidemic sizes and peaks. The future course of these epidemics and others around the world, of course, depends on the ability to maintain burdensome control measures over an extended period.

In several countries with high-performing healthcare systems where SARS-CoV-2 transmission has been established earlier, both supplies of personal protective equipment in hospitals and the availability of services has been problematic for COVID-19 care, and in all locations, ICU bed capacity is limited.^18^ Plans are urgently needed to mitigate the effect of COVID-19 outbreaks on the local healthcare system.

## Data Availability

Publically available data sources were used in this study. Raw data for this manuscript can be found at the github link provided.

https://github.com/c2-d2/COVID-19-wuhan-guangzhou-data

## Funding and acknowledgments

This work was in part supported by Award Number U54GM088558 from the National Institute Of General Medical Sciences. The content is solely the responsibility of the authors and does not necessarily represent the official views of the National Institute Of General Medical Science or the National Institutes of Health.

We thank the entire Center for Communicable Disease Dynamics team for providing a supportive environment for this work. RL thanks her fellow doctoral students for their support.

## Contributors

All authors contributed to the study design, RL analyzed the data and wrote the first draft of the manuscript, and all authors participated in the writing, reviewing, and editing of the manuscript.

